# A Serious Game (ACE of Hearts) to Support Mental Health among Young People with Adverse Childhood Experiences: A Mixed Methods Feasibility and Acceptability Evaluation

**DOI:** 10.64898/2026.07.21.26358490

**Authors:** Natalie Bisal, Haiou Zhu, Harsimran Sansoy, Isabelle Butcher, Minhua Ma, Kamaldeep Bhui

## Abstract

**Background:** Adverse childhood experiences (ACEs) have life-long detrimental effects on physical and mental health. ACE impacted young people are under-represented in research and cautious about seeking help. Practitioners fear re-traumatisation during assessment and treatment. Innovative approaches to care are needed. To respond, we co-designed a serious game called ‘ACE of Hearts’ (AoH) focusing on promoting awareness of ACEs and providing guidance and support. This paper reports on a feasibility and acceptability study and a process evaluation of mechanisms.

**Method:** A diverse cohort of young people aged between 12-24 years, reporting three or more ACEs, and living in different geographical areas of England were recruited through trusted partner organisations. Following informed consent, they were provided access to AoH for three months. Feasibility (rates of recruitment, uptake, engagement, retention, and follow-up), acceptability (affective attitude, burden, ethicality, intervention coherence, perceived effectiveness, self-efficacy), demographic and mental health outcome data were collected at baseline, 1- and 3-month follow-up. Process evaluation interviews at 3 months assessed mechanisms, acceptability and feasibility.

**Results:** Of 40 eligible subjects, 36 completed the baseline assessment and 22 downloaded AoH. Twenty participants completed the questionnaires at 1 and 3 months (91% follow-up of those accessing AoH); 19 participants completed all assessments (86%). Many participants valued the central cosy den space and its customisation options. Engagement rates for 4 mini-games ranged from 32% to 73%. Most participants enjoyed playing AoH, found it easy to use, and felt it helped them reflect on their experiences. The process evaluation found positive views of the game design, style, and content along with perceived benefits through education, awareness, emotional connection, and considering help-seeking. Several improvements for access, relevance, acceptability, engagement and retention were recommended. No adverse events were reported.

**Conclusions:** AoH was found to be feasible to use and acceptable and recommendations were given for improvements.

## Introduction

Mental health difficulties among young people are a growing concern, with up to three-quarters of mental illness emerging before the age of 25 years (1, 2). Despite this, many mental health problems remain unrecognised and untreated (3). Many young people do not access the necessary mental health support due to barriers such as long waiting times, not meeting clinical thresholds, lack of recognition and awareness in what help is available, and a fear of mental-health related stigma and discrimination (4–6).

Adverse childhood experiences (ACEs) include abuse, neglect, or household dysfunction that directly impacts the child and the safety of their environment (7–9). Young people exposed to childhood adversity face greater challenges and are more likely to experience poor health outcomes across the lifespan (9–12). Exposure to multiple ACEs is associated with a greater risk of poor physical and mental health, health-harming behaviours, reduced educational and employment outcomes, premature mortality (9, 10, 13–20), and substantial societal costs (10, 21, 22) estimated at £42 billion annually in the UK (21). Furthermore, young people affected by ACEs may struggle to narrate or make sense of their experiences, and recalling these events may be distressing, which can lead to avoidance of help-seeking and difficulties navigating complex health and social care systems. This highlights the need for innovative, accessible, and engaging approaches to mental health promotion and prevention tailored to young people affected by ACEs (23).

Digital mental health interventions offer a potentially promising, scalable route to early intervention for young people (24–27), including those affected by ACEs (28). They can help to address barriers related to stigma, privacy and anonymity, and reluctance or inability to access mental health services (26, 27). Many young people already use online tools (e.g., social media) to access informal support, seek social connection, and explore self-identity (29–31). Digitally delivered mental health resources therefore offer an appealing and acceptable approach (32) and represent a potentially cost-effective option (33).

There is increasing evidence for the value of serious games for mental health (34–37). Serious games are designed for educational, therapeutic and/or health improvement purposes (38, 39). They offer interactive experiences that leverage the appeal of digital opportunities for immersive play, exploring narratives, reward, and personalisation (34, 37, 40). The proposed mechanisms of benefit include distraction, engagement, psychoeducation, exposure-based desensitisation, cognitive restructuring via interactive problem-solving, emotional processing through narrative and metaphor, and social connectedness (36, 37, 40). Serious games can be self-guided and used in diverse settings, which may reduce stigma and widen reach among young people (41).

To our knowledge, no digital serious games have specifically targeted ACEs and mental health. To address this gap, we co-designed a serious game with key stakeholders as part of ATTUNE, funded by the UKRI programme Adolescent Mental Health and The Developing Mind. This study aimed to assess the feasibility and acceptability of a co-designed digital mental health serious game, *ACE of Hearts* (AoH), in young people, and understand potential mechanisms and further explore acceptability and feasibility via a process evaluation.

## Methods

### The serious game

AoH is a novel co-designed serious game combining playful interaction, psychoeducation, and health promotion, with opportunities for reflection on care and support options through recovery narratives. Metaphor and multimodal storytelling are central to the narratives and aim to offer playful and accessible engagement with difficult topics. The game comprises a central ‘cosy den’ space, which connects four mini-games, each focused on a different cluster of ACEs identified during preceding experience workshops as priority themes:

1. bereavement and caring responsibilities (*Horse and Foal*)
2. financial hardship (*Hard Times)*
3. disability (*Dial It Back*)
4. gender identity and dysphoria (*Out of My Shell*).

Players could sequentially unlock mini-games, and the game was designed for self-directed, independent use on smartphones and tablets. Figure 1 shows visual representations of AoH.

**Figure 1.**
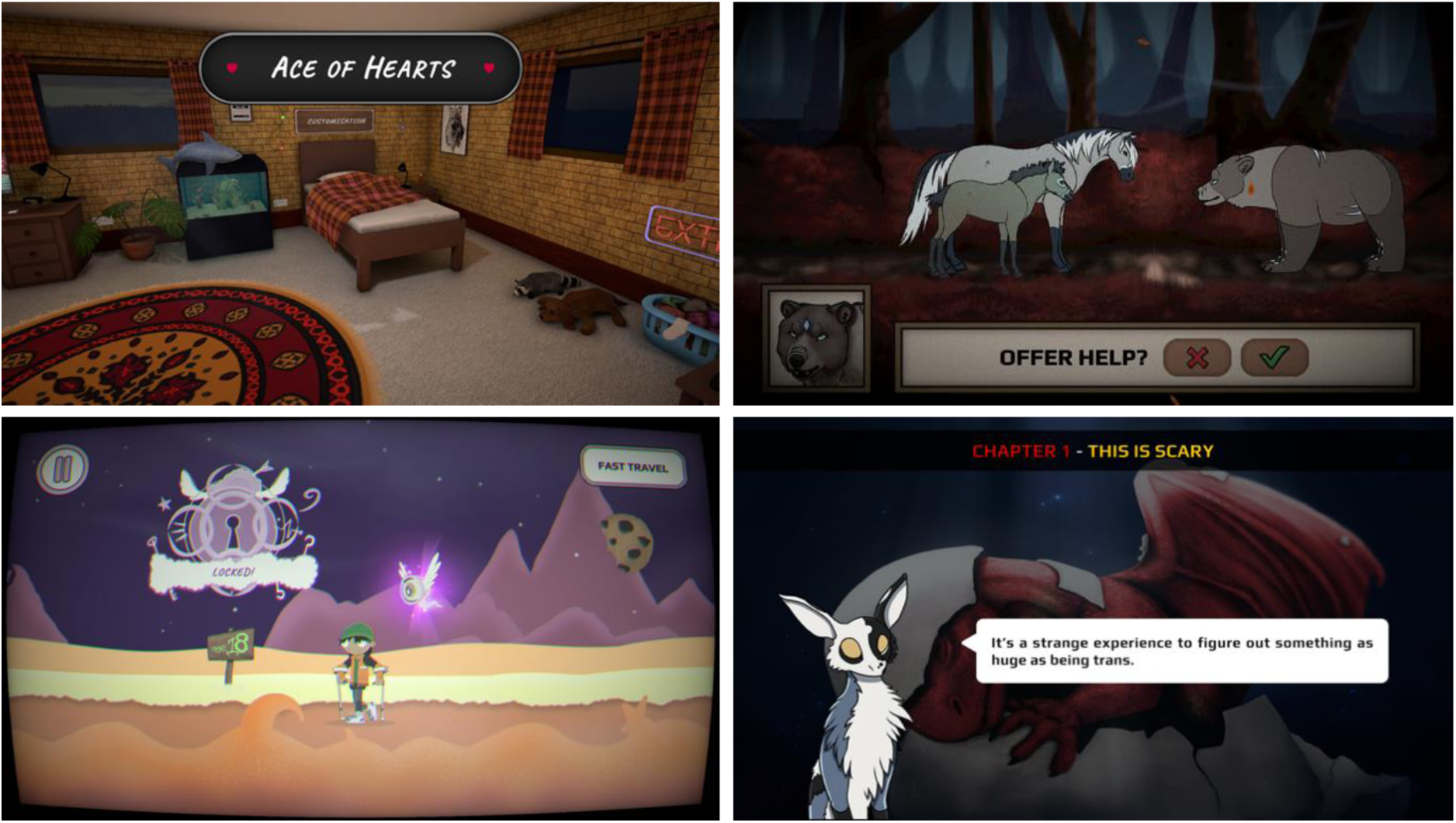
Visual representation of the serious game, displaying a screenshot from: (a) central *cosy den* hub (*top left*), (b) Horse and Foal (*top right*), (c) Dial It Back (*bottom left*), and (d) Out of My Shell (*bottom right)* mini-games.

AoH was developed using an iterative, experience-based co-design approach integrated with agile game development (42), enabling rapid iteration based on participant feedback (43) while engaging end-users and stakeholders as equal partners to reflect lived experience (44) and enhance acceptability (45, 46). Twelve workshops across eight co-design sprints were conducted with 18 diverse lived experience young people, alongside five adult stakeholders and a multidisciplinary game development team (47). See (42) for a detailed description of the game design, content, and development processes.

### Study design

We conducted a mixed-methods feasibility and acceptability study. Data were collected at baseline, one-month and three-month follow-up using online questionnaires. Feasibility and acceptability outcomes assessed rates of recruitment, uptake, engagement, retention, follow-up, and acceptability. We also collected qualitative process evaluation data via one-to-one semi-structured interviews at three-months.

### Setting and participants

This study utilised a digital platform for online recruitment and evaluation. Eligible participants were young people aged between 12-24 years, who had experienced three or more ACEs (using the ACE-IQ (48)), had access to a smartphone or tablet with internet connection, were affiliated with a partner organisation in England, and were able to read and understand English. Individuals were excluded if they had previously been involved in the game development phase.

### Procedure

Young people were recruited (August 2024-May 2025) through community organisations, educational settings, relevant websites and newsletters, and face-to-face sessions. Young people expressed interest and were provided with an information sheet prior to giving informed consent, or assent and parental consent for participants under 16 years. Additional support was available during consent and eligibility assessment. Eligible participants completed an online baseline assessment with a researcher, then received instructions to download the game, with support and reminders available. Participants were given access to AoH for a period of three months. Participants were reimbursed for their time and contribution to the study at baseline (£10 voucher) and three months (£15 voucher). Safeguarding procedures were in place throughout the study. Affiliated organisations were notified if participants met clinical thresholds for depression or anxiety (PHQ-9 ≥ 12, GAD-7 ≥ 12, or endorsement of suicidal ideation) at any assessment point.

NHS Research Ethics Committee approvals were granted (NHS REC number: 23/WM/0105) with sponsorship from the University of Oxford.

### Measures

#### Feasibility and acceptability measures

We recorded expressions of interest, assents or consents, eligibility, uptake (number of young people who accessed AoH), completed assessments at baseline, one and three-month follow-up, and serious game engagement (in-game data on number of mini-games played).

Acceptability was assessed using a 6-domain measure (e.g., burden, self-efficacy; adapted from Acceptability of Health Apps among Adolescents (49)), with full details provided in Supplementary Material 1.

#### Sociodemographic variables

At baseline, participants self-reported their age, gender, ethnic background, sexual orientation, trans status, religion, neurodevelopmental conditions, whether they were born in the UK, geographical location, education and employment status, parent/caregiver status, whether their parents/caregiver were born in UK, and main parent/caregiver education and employment status.

#### Mental health

Participants completed the following validated measures at baseline, one and three-month follow-up: depression (PHQ-9 (50, 51)), anxiety (GAD-7 (52)), mental wellbeing (SWEMWBS (53)), post-traumatic stress (CRIES (54)) and emotion regulation (DERS-SF (55) (see Supplementary Material 1).

### Qualitative process evaluation

Semi-structured interviews were conducted online (using Microsoft Teams) or via written responses with participants who had accessed AoH. Interviews sought to explore participants’ experience of AoH using 16 open-ended questions (Supplementary Material 2). Interviews were documented using researchers’ notes to minimise participant burden and discomfort when discussing potentially sensitive experiences. Participants were invited to review and verify the recorded notes prior to analysis and renumerated (£10 voucher) for their participation.

### Data Analyses

Descriptive statistics summarise demographic characteristics, mental health outcomes, feasibility and acceptability (using frequencies and percentages for categorical data and means and standard deviations for continuous data). Pre-post outcomes were calculated as mean differences with 95% confidence intervals across baseline, one-month, three-month follow-up assessments for participants who completed all study assessments. Statistical analyses were conducted using SPSS (version 30.0).

Qualitative data were analysed using an inductive thematic analysis approach (56, 57), including; familiarisation, generating initial codes, searching for themes, collectively reviewing themes, and defining and naming the themes. NB coded all data and HZ, IB coded 75% of the data. Codes and themes were refined, and discrepancies were resolved by reaching consensus. Qualitative analyses were conducted in Microsoft Excel, providing a transparent audit trail.

## Results

### Feasibility

Feasibility outcomes are summarised in Table 1 and participant flow is shown in Figure 2. A total of 36 young people completed baseline, of which 22 accessed AoH at least once. Reasons for a lack of uptake included difficulties with access or forgetting to download the game. Engagement levels decreased with each successive mini-game, with the proportion of mini-games played ranging from 73% (*Horse and Foal*) to 32% (*Out of My Shell*). Follow-up assessment completions were high at all timepoints.

**Table 1.** Feasibility outcomes of ACE of Hearts.

| Outcomes | Participants |
| --- | --- |
| Number of young people expressed interest in the study, n | 78 |
| Rates of young person consent and assent, n (%) | 48 (61.5) |
| Rates of eligibility | 40 (83.3) |
| Rates of enrolment (baseline assessment completed), n (%) | 36 (90.0) |
| Rate of uptake (serious game accessed), n (%) | 22 (61.1) |
| Rates of assessment completion, n (%) |  |
| One-month follow-up | 20/22 (90.9) |
| Three-month follow-up | 20/22 (90.9) |
| Rate of retention (all assessments completed), n (%) | 19/22 (86.4) |
| Rates of engagement (mini-games played), n (%) |  |
| Mini-game 1: Horse and foal | 16/22 (72.7) |
| Mini-game 2: Hard times | 12/22 (54.5) |
| Mini-game 3: Dial it back | 10/22 (45.5) |
| Mini-game 4: Out of my shell | 7/22 (31.8) |

**Figure 2.**
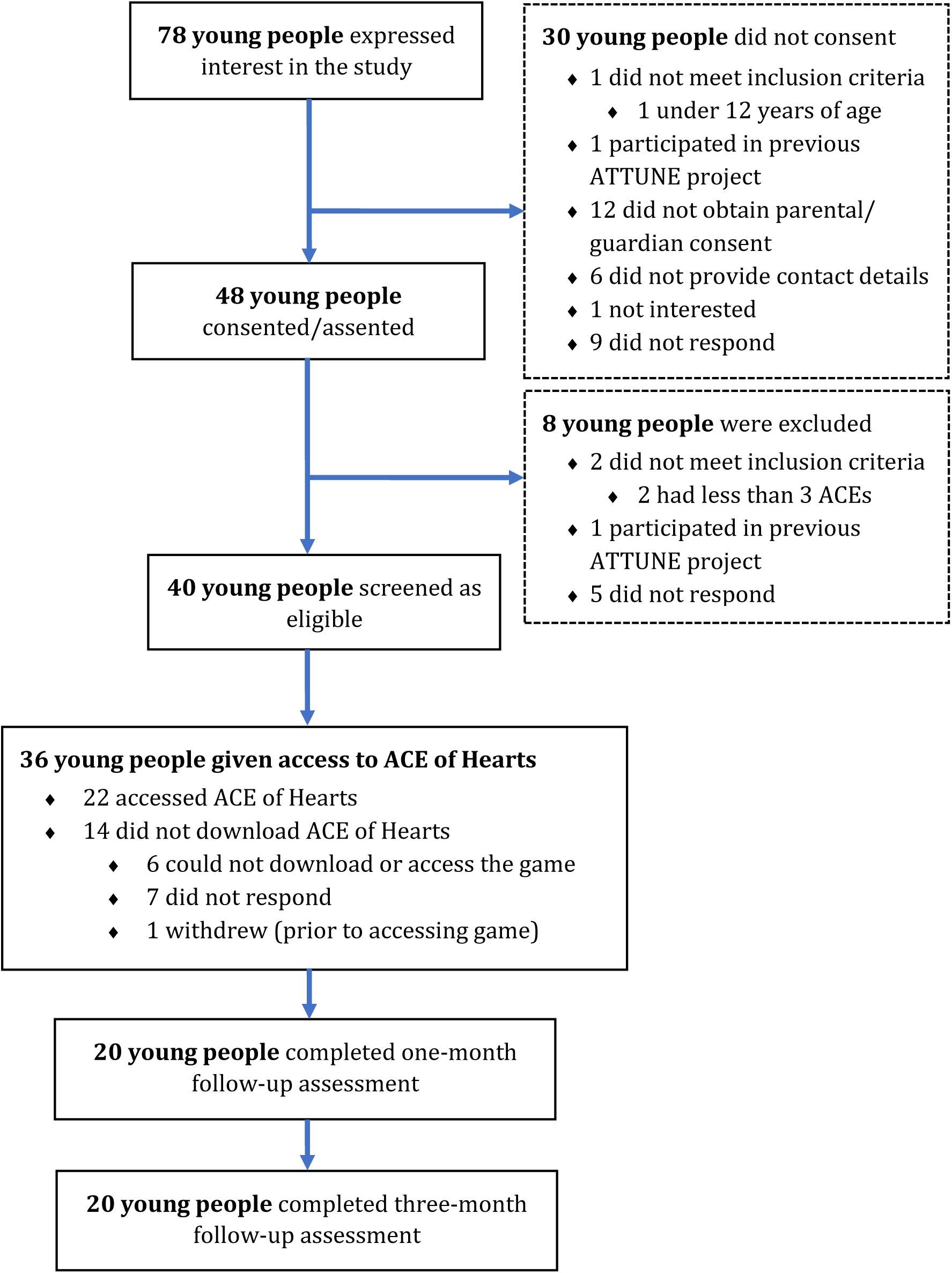
Participant flowchart diagram (based on CONSORT; (58)).

### Participant characteristics

Participant baseline characteristics and PHQ-9 and GAD-7 scores are summarised in Table 2 (additional characteristics are provided in Supplementary Material 3).

**Table 2.**
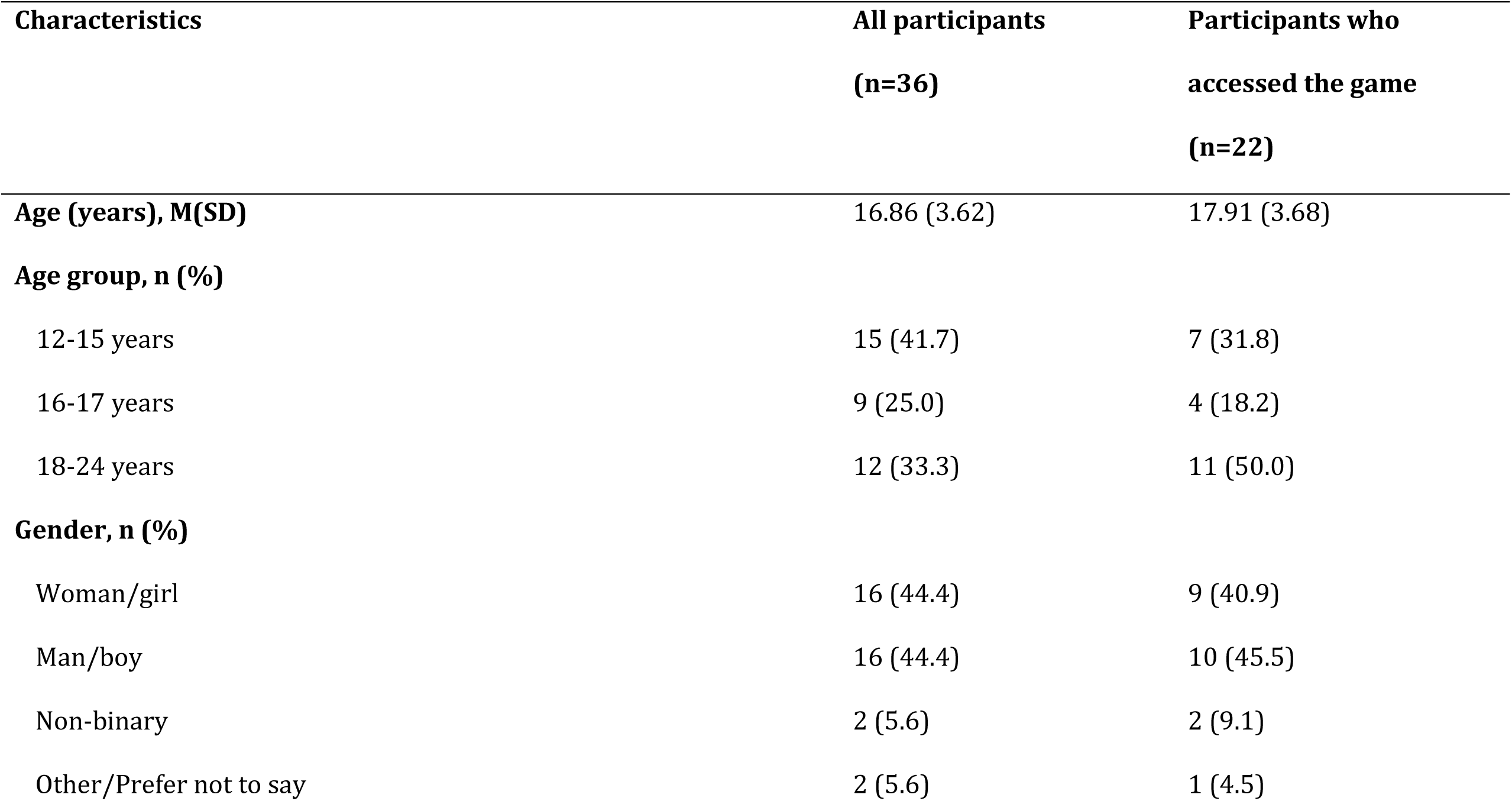
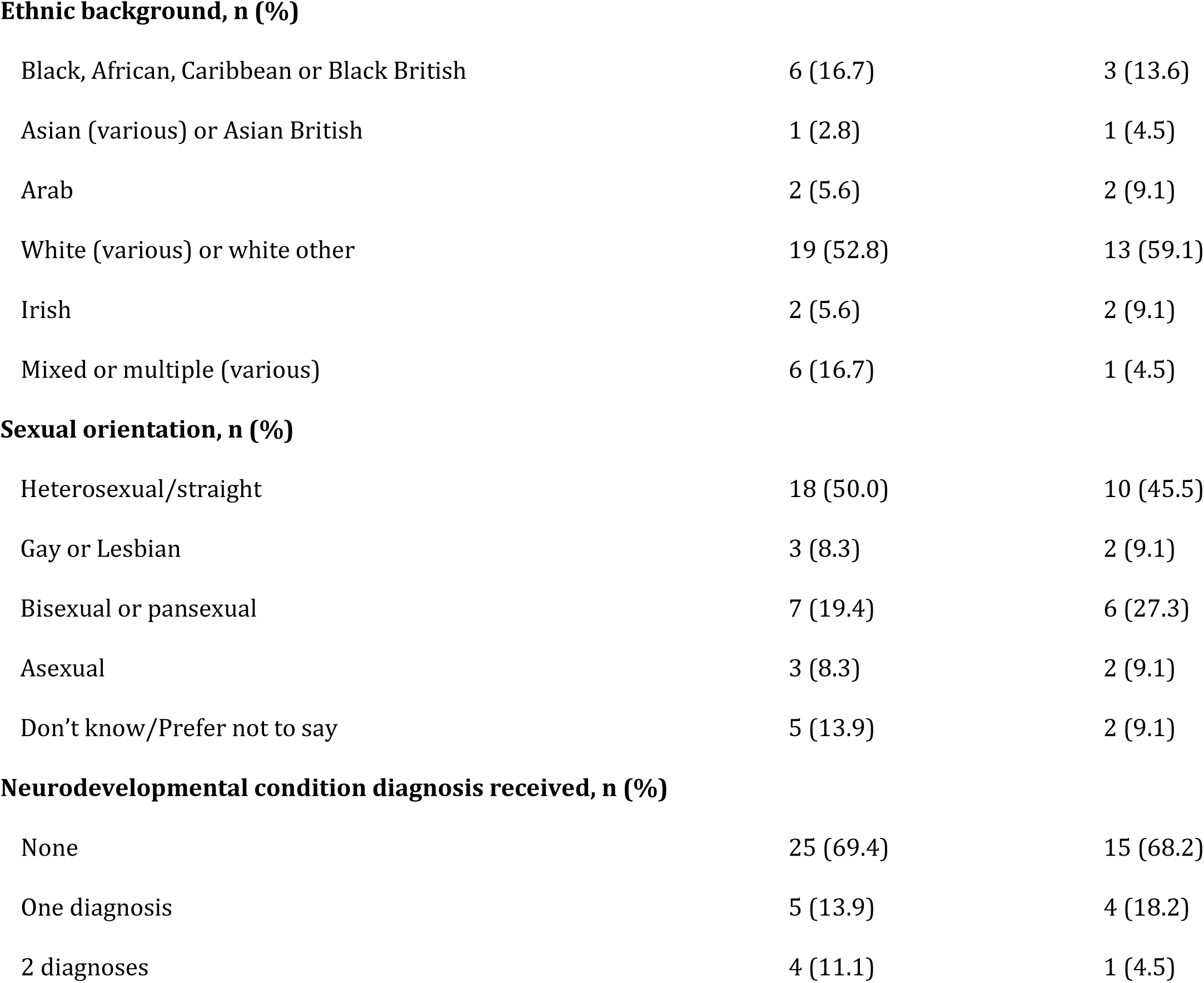
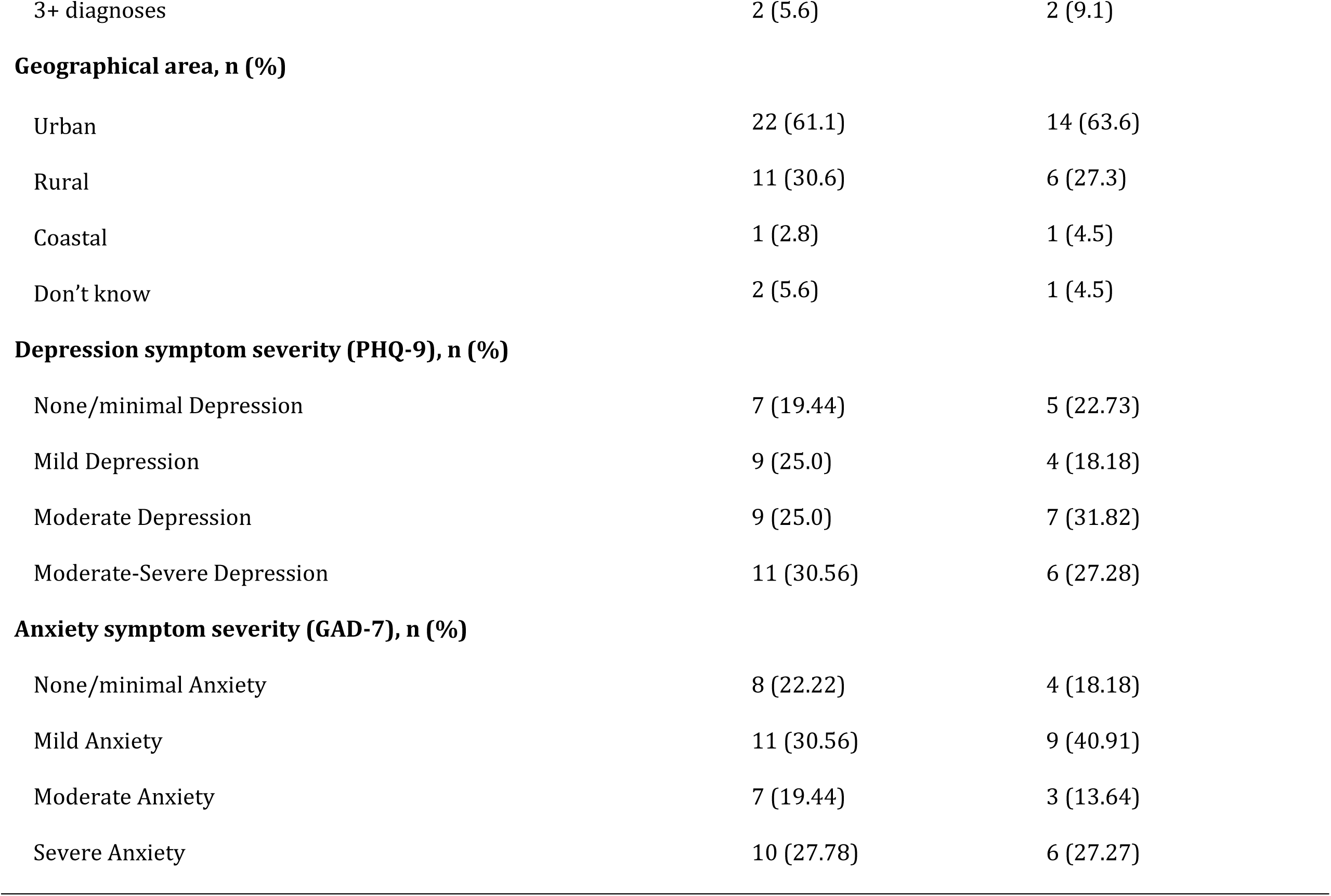
Characteristics of the sample by all participants who completed the baseline assessment (n=36) and participants who accessed the serious game (n=22)

| Characteristics | All participants<br>(n=36) | Participants who<br>accessed the game<br>(n=22) |
| --- | --- | --- |
| Age (years), M(SD) | 16.86 (3.62) | 17.91 (3.68) |
| Age group, n (%) |  |  |
| 12-15 years | 15 (41.7) | 7 (31.8) |
| 16-17 years | 9 (25.0) | 4 (18.2) |
| 18-24 years | 12 (33.3) | 11 (50.0) |
| Gender, n (%) |  |  |
| Woman/girl | 16 (44.4) | 9 (40.9) |
| Man/boy | 16 (44.4) | 10 (45.5) |
| Non-binary | 2 (5.6) | 2 (9.1) |
| Other/Prefer not to say | 2 (5.6) | 1 (4.5) |
| **Ethnic background, n (%)** |  |  |
| Black, African, Caribbean or Black British | 6 (16.7) | 3 (13.6) |
| Asian (various) or Asian British | 1 (2.8) | 1 (4.5) |
| Arab | 2 (5.6) | 2 (9.1) |
| White (various) or white other | 19 (52.8) | 13 (59.1) |
| Irish | 2 (5.6) | 2 (9.1) |
| Mixed or multiple (various) | 6 (16.7) | 1 (4.5) |
| **Sexual orientation, n (%)** |  |  |
| Heterosexual/straight | 18 (50.0) | 10 (45.5) |
| Gay or Lesbian | 3 (8.3) | 2 (9.1) |
| Bisexual or pansexual | 7 (19.4) | 6 (27.3) |
| Asexual | 3 (8.3) | 2 (9.1) |
| Don't know/Prefer not to say | 5 (13.9) | 2 (9.1) |
| **Neurodevelopmental condition diagnosis received, n (%)** |  |  |
| None | 25 (69.4) | 15 (68.2) |
| One diagnosis | 5 (13.9) | 4 (18.2) |
| 2 diagnoses | 4 (11.1) | 1 (4.5) |
| 3+ diagnoses | 2 (5.6) | 2 (9.1) |
| **Geographical area, n (%)** |  |  |
| Urban | 22 (61.1) | 14 (63.6) |
| Rural | 11 (30.6) | 6 (27.3) |
| Coastal | 1 (2.8) | 1 (4.5) |
| Don't know | 2 (5.6) | 1 (4.5) |
| **Depression symptom severity (PHQ-9), n (%)** |  |  |
| None/minimal Depression | 7 (19.44) | 5 (22.73) |
| Mild Depression | 9 (25.0) | 4 (18.18) |
| Moderate Depression | 9 (25.0) | 7 (31.82) |
| Moderate-Severe Depression | 11 (30.56) | 6 (27.28) |
| **Anxiety symptom severity (GAD-7), n (%)** |  |  |
| None/minimal Anxiety | 8 (22.22) | 4 (18.18) |
| Mild Anxiety | 11 (30.56) | 9 (40.91) |
| Moderate Anxiety | 7 (19.44) | 3 (13.64) |
| Severe Anxiety | 10 (27.78) | 6 (27.27) |

Participants who played AoH generally presented similar characteristics as those who did not access the game, though some small differences were observed on visual inspection of the data. A lower proportion of participants who played the game had experience of foster care (0% versus 11%) and were in school/college (50% versus 61%) compared to the total sample, respectively. In contrast, participants who played the game were more likely to identify as bisexual or pansexual (27% versus 19%) and be employed (55% versus 36%) compared with the total sample. Depressive and anxiety symptoms at clinical thresholds were similar by games access.

### Acceptability

Among participants who accessed AoH, 20 (out of 22; 91%) completed the acceptability questionnaire (see Supplementary Material 4). Most participants reported that they enjoyed using AoH (85%), found it simple (70%) and easy to use (85%), and felt it could help them to think about their own life experiences (80%) and acknowledge difficult experiences they had been through (80%). Just over half of participants (55%) understood how to use the game features, while 65% reported feeling confident using AoH without reminders.

### Mental health outcome measures

Supplementary Material 5 shows the pre-post scores for the mental health outcome measures at baseline, one- and three-month follow-up. Although this study was not designed to assess the efficacy of the serious game, no harms or unintended outcomes were reported. While not suitable for formal statistical testing, the observed patterns provided preliminary indications of positive trends in anxiety, depression, and mental wellbeing, and importantly no worsening.

### Qualitative process evaluation of the serious game

Fifteen participants (75%; of 20 invited) completed an interview. Most were aged 18-24 years (60%), from a White background (73%), and identified as female (40%), male (40%) or non-binary (13%) (see Supplementary Material 6). Thirteen interviews were conducted online with a researcher and two were completed independently by participants in written format. Eight (67%, out of 12 invited) participants verified that their data were accurate and complete. Analysis resulted in four overarching themes: (1) Design features, style and customisability, (2) User experience and accessibility, (3) Characters, stories and content, and (4) Perceived impact. Illustrative quotes and researcher notes are presented in Table 3.

**Table 3.** Themes, subthemes, and illustrative participant quotes or researcher notes.

| Themes and subthemes | Quote / note No. | Quote or note examples |
| --- | --- | --- |
| **Key theme 1: Design features, style, and customisability** |  |  |
| Overall design, feel and appearance | 1 | *"This is one of the most unique games I've ever played... There is nothing currently or previously out there that accurately represents young people who have caring responsibilities and so this is a great tool for lots of young people."* (P08; researcher note) |
|  | 2 | *"Making it visually nice to play makes people engaged to play from the beginning if people aren't motivated to play. The first and third game, I really like the art style and game design."* (P04; researcher note) |
|  | 3 | *"Some of the games just felt a bit simple and slow, I found them too easy or wanted them to go faster as the speed was making me a little bit bored at times... I didn't really like the way the game worked, it felt very repetitive and long, it nearly put me off the game as a whole but I got through it and moved on to more enjoyable mini-games."* (P03; quote) |
| Interactive elements and customisation | 4 | *"Loved the cosy room and the ability to customise this in one's own way and the fact that one could change little things and personalise this was great e.g. the guitar, lots to do in this room and was/is a chill out space after playing the game."* (P09; researcher note) |
|  | 5 | *"...a little journal type area where you could type away and life experience e.g. like the notes app on the phone. And the game could store these... Meditating space where it says 321 breath in and breath out – so a practical place and tips." (P05; quote)* |
| **Key theme 2: User experience and accessibility** |  |  |
| User experience, navigation and ease of use | 6 | *"It could be a bit confusing when you were in a room to explore but didn't know where to start and what to do. For people unfamiliar with such game formats, it could be confusing. May be more signposting and direct guidance to play the game. It would also be beneficial for tailoring for people, e.g., neurodiverse users." (P06; researcher note)* |
|  | 7 | *"I don't know how it could be more engaging. If you download a game, most of the time they send you notifications or something 'hey you haven't looked at me for two days' or something, if the person who saw the notification had a free moment, it would prompt them to play." (P11; researcher note)* |
|  | 8 | *"I liked the simplicity of most of the games as well... It [was] very mostly very low effort, most of the games were very easy and I could easily play them while watching tv or talking to someone." (P03; quote)* |
| Accessibility of the mobile game | 9 | *"Yes I thought it was [accessible] apart from Mina's story because I found the controls didn't work very well with my dyspraxia. Other than that it was fine though because I don't have many other accessibility needs." (P03; quote)* |
|  | 10 | *"Maybe, it might be better if it was a desk top game. That's a space I dedicated to play the game. It's quite easy for me to pick up the phone but also easy to distract by other thing using my phone" (P12; researcher note)* |
| **Key theme 3: Characters, stories and content** |  |  |
| Appropriateness and relatability | 11 | *"I liked that multiple of the characters represented experiences I've been through and I saw myself in them, I've never had that with a game before, although I don't play video games very often." (P03; quote)* |
|  | 12 | *"In the Horse and Foal game the anxious aspect [referring to the anxiety bar] resonated with my experience and reminded me it's only temporary and doesn't last forever." (P02; researcher note)* |
|  | 13 | *"Some of it was relatable but not a lot of it which is good thing because if it was all relatable that would be a bummer. When I reflect on it I'm not going to complain about not being able to relate to negative life experiences." (P07; researcher note)* |
| Intended for a younger audience | 14 | *"... but maybe I was a bit too old for them. I know I was at the upper end of the target demographic so I noticed it was targeted to people younger than me in the way it was portrayed. The first story was horse-riding and there was a metaphor for being a horse and that feels a bit childish.. I think it depends on the target audience. I'm 25 now so at that age of the end group it would make sense to have more complex characters." (P11; researcher note)* |
|  | 15 | *"Loads of outcomes for people in the ACEs categories (smoking, alcoholism, earlier death), they are real dangerous risks, kids who have been abused, self-harming activities (smoking, drinking, drugs), that the game doesn't explore which makes sense not including as not wanting to talk about them with younger kids... If targeted at older audience, those kinds of things definitely need to be brought up." (P04; researcher note)* |
| Storytelling, metaphors and arts-based methods | 16 | *"I loved, the gender dysphoria, it might have been my favourite of the bunch. I loved the egg analogy; you hear about that all the time in the trans community. It's really cool that the term was incorporated into the game, in a youth friendly way. Can definitely see it being easy to understand for teenagers/pre-teens." (P04; researcher note)* |
|  | 17 | *"Caring for parent(s) as a young age can bring shame and this was portrayed well in the game and was not super super judgemental." (P08; researcher note)* |
|  | 18 | *"I really like the setting and customisation, e.g., painting, guitar. Take away the heaviness of the game, and be able to interact with the game and make fun" (P12; researcher note)* |
| **Key theme 4: Perceived impact** |  |  |
| Education, awareness and reflection | 19 | *"I would say a bit of yes, especially in raising awareness. For someone who hasn't thought about these issues before, it could be quite eye-opening and change their mindset." (P06; researcher note)* |
|  | 20 | *"Thoughtful. It made me think more about it, the situation. Even if you haven't experienced those it makes you think about your own experiences even if it's not the same but similar."* (P11; researcher note) |
|  | 21 | *"...a lot of the experiences weren't my experiences but they were interesting and it can be interesting to know about the experiences about other people, I know the game was to help heal people from these experiences but it was cool to learn about others experiences."* (P07; researcher note) |
| Emotional impact and connection | 22 | *"I think it reassured me that these experiences are shared by a lot of people as well as fictional characters and it did help me reflect a bit."* (P03; quote) |
|  | 23 | *"It [the game] has helped in their journey and the shame that they experienced in their own journey in life"* (P08; researcher note) |
|  | 24 | *"They covered really dark topics and part of you feels sad when you read it, empathy for the characters, but you also feel seen and not normalises your experience but gives it a shape, a name."* (P04; researcher note) |
| Psychological safety | 25 | *"Sometimes, it makes you feel emotional and stressful. When the darkness chases and approaches [horse and foal mini-game], it could be a bit stressful. For people with severe mental health challenges, that could be triggering ... Personally, I was okay. It's more about raising awareness and for advocating for these kinds of ACEs."* (P06; researcher note) |
|  | 26 | *"...everyone is different so if someone is really currently traumatised this game could be triggering but then trigger warnings could be used perhaps."* (P08; researcher note) |
| Help-seeking | 27 | *"Yes, it encouraged me to consider seeking help. It gave me the mindset that maybe I do need to take action, and made me feel that talking to a counsellor is a good decision." (P06; researcher note)* |
|  | 28 | *"Good to start a conversation, maybe with teens who might not be willing to open up and talk about it or early 20s when you've matured more, when it would be easier to talk to someone about it, without having to go to someone about it." (P11; researcher note)* |
|  | 29 | *"It could do a better job of connecting people to outside sites (Samaritans, suicide hotline etc) people you can talk to if you need to talk to...Especially if you come out of one of the minigames, and you're feeling down you can click one of the icons and see if you can talk to someone there." (P10; researcher note)* |

#### 1. Design features, style, and customisability

##### Overall design, feel and appearance

Participants suggested the game was fun, unique, and helpful to consider mental health and ACEs (*researcher note 1*). They generally liked the visual design, arts content, and interactive elements (*researcher note 2*). However, for some people, the mini-games (e.g. *Horse and Foal)*, were considered too lengthy and slow, leading to drifting attention or boredom (*quote 3*). Others enjoyed the slower pace, as calmer and more relaxing than ‘overstimulating’ games.

##### Interactive elements and customisation

Many participants liked the personalisation features, particularly the *cosy den* (*researcher note 4*). They suggested ways to support engagement and reduce cognitive effort (e.g., through puzzles), expand the range of topics covered, and include guided activities (e.g., journaling, breathing, meditation) (*quote 5*). Some participants said they had limited control over predetermined storylines. Allowing choice over which mini-game to play first might tailor experiences.

#### 2. User experience and accessibility

##### User experience, navigation and ease of use

Several participants wanted clearer signposting and guidance (*researcher note 6*), and notifications to prompt engagement (*researcher note 7*). Nonetheless, many found AoH simple, requiring minimal effort to use (*quote 8*).

##### Accessibility

AoH was generally considered accessible, convenient, and useful for engaging young people. However, some participants experienced difficulties navigating the controls which may present challenges for some users (including neurodivergent young people; *quote 9*). A desktop-based version of the game may increase accessibility and ease of use and reduce distractions (*researcher note 10)*.

#### 3. Characters, stories and content

##### Appropriateness and relatability

Several participants were able to relate to the characters and stories depicted, with some stories reflecting their own experiences (*quote 12)*. The topics presented were ‘*real issues’* faced by young people and perceived as relevant to ACEs (*researcher note 12)*. For others, the content did not fully reflect personal experiences, which was both beneficial and limiting. Contrasting content helpfully allowed emotional distance (*researcher note 13)*, while storylines lacking resonance or the ‘wrong message’ were less helpful. Additional mini-games on other topics, such as social media, screen use, school and exam stress, friendships, peer pressure, and family difficulties, were proposed.

##### Intended for a younger audience

It was widely felt that AoH was suited to a younger audience (e.g., teenagers), owing to the game style, characters, and topics covered (*researcher note 14)*. For older groups (particularly over 18 years), the game was viewed as less relevant. More age-appropriate topics (e.g., addiction, substance use), characters and storylines with greater complexity were recommended (*researcher note 15)*.

##### Storytelling, metaphors and arts-based methods

Many participants were excited about the topics and found metaphors, storytelling, and creative arts helpful for delivering challenging content (*researcher note 16*). They valued the role of interactive play to learn about mental health and ACEs. Participants valued the youth-friendly, non-judgemental approach (*researcher note 17*) and the balance between fun and calming features (e.g., the *cosy den)* and more serious, emotionally heavy elements (*researcher note 18*).

#### 4. Perceived impact

##### Education, awareness and reflection

AoH raised awareness, helping participants learn about mental health and ACEs (*researcher note 19)*. It supported some participants to reflect on their experiences (*researcher note 20)*. Many reported a better understanding of others’ life experiences, and that AoH might educate people about issues such as gender dysphoria (*researcher note 21)*.

##### Emotional impact and connection

Participants said their experiences were represented and they felt seen and understood. They valued validation of different emotions, reassured by others sharing similar experiences (*researcher notes 22-23)*. The stories could also evoke emotions such as empathy and sadness, which, fostered a sense of connection with the characters and their narratives (*researcher note 24)*. Others reported feeling comforted, hopeful, or happy when positive outcomes occurred for the characters.

##### Psychological safety

Most participants reported feeling safe while playing the game. However, several noted content which could be challenging and potentially triggering (*researcher note 25)*. Participants recommended more trigger warnings (*researcher note 26),* with pause options or return to the cosy den option.

##### Help-seeking

Some said the game could encourage help-seeking (*researcher note 27)* or initiate conversations about mental health (*researcher note 28)*. However, many did not seek help themselves, either because they did not need support or had already received it. Participants noted a need for clearer signposting and interactive formats, such as clickable links to external websites or videos to enhance engagement and seeking further support (*researcher note 29)*.

## Discussion

This study overall finds high rates of acceptability, retention, and outcome completion at one and three months. Uptake and engagement rates need further improvement.

### Interpretation in context

Incomplete downloads and usage of AoH is consistent with broader evidence for digital mental health interventions among young people (32), particularly for unguided and self-directed resources (59–61). Incorporating prompts or reminders and offering optional tailored support may improve motivation for individuals preferring more structure (62). In addition, personalisation, appealing design, and content tailored to individual needs, including greater user choice over game elements and sequencing, might help sustain engagement and enhance user experience (23, 26, 63, 64).

In terms of uptake, 14 out of 36 participants did not download or were unable to access AoH (e.g., due to device) despite reminders and offers of real-time support. People from socioeconomically disadvantaged backgrounds are more likely to face barriers accessing mental health interventions (65). Alternative options are therefore needed to increase access for digitally marginalised young people such as a browser-based desktop version. Future, larger-scale studies with more diverse samples should investigate how individual and contextual factors influence engagement for youth mental health.

Age appropriateness and perceived relevance are critical for acceptability (63). Although individuals aged 18–24 years are often classified within extended definitions of adolescence, their cognitive development, life transitions, and mental health needs differ from those of younger adolescents (66, 67). Our findings support greater age-stratified design and tailoring of content, narrative complexity, and gameplay mechanics.

Recognition of shared experiences supported validation, emotional engagement, and self-reflection (68, 69), consistent with psychological processes of bonding social capital (70). Of interest, engagement with narratives dissimilar to one’s own appeared to encourage empathy, openness, and perspective-taking (71–73). These processes underpin bridging social capital (70). Further research should examine these proposed mechanisms and their relevance to youth mental health outcomes.

Findings on storytelling and metaphor resonate with therapeutic literature highlighting the value of emotional distance in promoting a sense of safety (74, 75). Video games are particularly suited to conveying metaphor because they operate across multiple channels, visual, auditory, haptic and interactive, creating an “ecosystem of meaning-making” (76). In AoH, multimodal metaphors were integrated throughout gameplay which may provide players with embodied experiences that extend beyond those typically achievable through traditional approaches alone (76, 77). These findings align with wider evidence that creative and digital interventions can promote understanding the mental health consequences of ACEs (78).

### Strengths and limitations

Given the limited research specifically focusing on ACE-related experiences among young people, this study contributes feasibility and acceptability data for an innovative co-designed serious game. The mixed-methods design provided rich insights into participants’ experiences (79). A high proportion of participants who accessed the game, played and completed outcomes, and participated in the process evaluation. Additionally, we actively recruited participants from diverse groups to enhance inclusion (23, 80) and made efforts to address the gender imbalance commonly observed in similar studies (23).

This study has several limitations. Retrospective qualitative interviews may have influenced participants’ ability to recall their experiences. Process evaluation relied on researcher notes, although participants were asked to verify recorded data prior to analysis. Participants who did not access the serious game were not followed up, limiting insights into the barriers to uptake.

### Implications

Overall, our findings align with literature suggesting that serious games may offer a feasible and engaging approach to supporting youth mental health (36, 81–84). Further co-design and iterative refinement is recommended to enhance acceptability and optimisation of the game. Future research is required on longer-term cost-effectiveness and implementation.

### Conclusion

Young people with ACEs viewed *ACE of Hearts* as acceptable, appropriate, and novel, with good retention at one and three months among those who accessed the game. Identifying barriers and facilitators to access, download and play to optimise impact in diverse populations will inform a future trial.

## Preprint note

This manuscript is a preprint and has not yet undergone peer review. Findings should therefore be considered as preliminary.

## Acknowledgements

The authors wish to thank the participants who generously gave their time to take part in this research as well as the wider ATTUNE team. We gratefully acknowledge the young people who co-designed *ACE of Hearts* and the YPAGs, whose insight and support were integral to the development of this work. We also wish to express our gratitude to the game development team, Graham Smith, Flick Broadley, and Jamie Jones, for their creativity, commitment, and significant contribution to the design and development of *ACE of Hearts*.

## Author contributions

**NB** made substantial contributions to the design of the work as well as recruitment, data collection, analysis and interpretation; drafted and substantially revised the work; approved submitted version.

**HZ, IB, HS** acted as researchers and made substantial contributions to the design of the work, as well as recruitment, data collection, and analysis; approved submitted version.

**MM** is the co-PI, made substantial contributions to the conception and design of the work, as well as acquisition, analysis and interpretation of data; co-applicant on the funding application; approved submitted version; led the work package that these data and outputs relate to.

**KB** is the PI, draw in funding, ethics review, data protection and safeguarding procedures, and made substantial contributions to the conception and design of the work, as well as acquisition, analysis and interpretation of data; substantially revised the work; approved submitted version; co-led the work package that these data and outputs relate to.

KB and MM act as guarantors, all authors accepting responsibility for all aspects of the study.

## Data availability statement

Research data are not shared, given the small sample sizes and detail on numerous intersectional characteristics. The data are not appropriate for complex secondary analyses. The research team welcome collaborations to develop more robust data sets.

