## Supplementary Materials for "A Serious Game (ACE of Hearts) to Support Mental Health among Young People with Adverse Childhood Experiences: A Mixed Methods Feasibility and Acceptability Evaluation"

**Supplementary Material 1.**

**eTable 1.** Description of acceptability and mental health outcome measures collected

| Outcome | Questionnaire | Description |
|---|---|---|
| Acceptability | Acceptability of Health Apps among Adolescents (AHAA) (49) | Acceptability was measured at one-month using 22 questions adapted from the AHAA. Participants responded to '*Yes*' and '*No*' response options on items measuring acceptability across 6 domains: *Affective Attitude*, *Burden*, *Ethicality*, *Intervention Coherence*, *Perceived Effectiveness*, and *Self-Efficacy*. |
| Depression | Patient Health Questionnaire (PHQ-9) (50, 51) | The PHQ-9 comprises 9-items measured on a scale from 0 (*not at all*) to 3 (*nearly every day*). Total scores range from 0 to 27 and higher scores indicate greater levels of depression. Scores were categorised into none to minimal (0-4), mild (5-9), moderate (10-14), moderately severe (15-19), and severe (20-27), with clinical threshold for anxiety of a score of 10 or above (44). The scale has good reliability and validity and has been used in culturally diverse populations. The measure showed good internal consistency for the total sample at baseline ($\alpha$ = .89). |
| Anxiety | Generalised Anxiety Disorder Assessment (GAD-7) (52) | The GAD-7 contains 7-items measured on a scale from 0 (*not at all*) to 3 (*nearly every day*). Total scores range from 0 to 21 and higher scores indicate greater levels of anxiety. Scores were categorised into minimal (0-4), mild (5-9), moderate (10-14), and severe (15-21), with clinical threshold for anxiety of a score of 10 or above (46). The measure showed good internal consistency for the total sample at baseline ($\alpha$ = .86). |
| Mental wellbeing | Short Warwick-Edinburgh Mental Wellbeing Scale (SWEMWBS) (53) | The SWEMWBS is a validated 7-item scale that measures predominantly functioning aspects of mental wellbeing. Items are assessed on a scale from 1 (*none of the time*) to 5 (*all of the time*). Total scores range from 7 to 35 and higher scores represent greater levels of mental wellbeing. The measure showed good internal consistency for the total sample at baseline ($\alpha$ = .82). |
| PTSD symptoms | Children's Revised Impact of Event Scale (CRIES) (54) | The CRIES is a validated 8-item scale that covers both intrusion and avoidance. Total scores range from 0 to 40 and a higher score indicates greater post-traumatic stress (PTSD) symptoms. |

| | | |
|---|---|---|
| | | The total mean was calculated total impact of events score along with the two subscales. The measure showed good internal consistency for the total sample at baseline (α = .89). |
| Emotion regulation | Difficulties in Emotion Regulation Scale short-form (DERS-SF) (55) | Participants aged 18 years or over completed the DERS-SF which is an 18-item scale comprising six subscales of strategies, non-acceptance, impulse, goals, awareness, and clarity. DERS-SF is measured on a 5-point Likert scale from 1 (*almost never*) to 5 (*almost always*) and total scores range from 18 to 90. Higher scores indicate greater difficulties with emotion regulation. The total mean was calculated total emotion regulation score along with each of the six subscales. The measure showed acceptable reliability for the total sample at baseline (α = .74). |

**Supplementary Material 2.**

Qualitative interview guide

1. How did you use the game? (e.g., where, time of the day, effort, frequency (every day, a few times a week, once a week))
2. How was your experience of playing it?
3. Was the game accessible to you? Did you have difficulties finding, opening, understanding what you are meant to do etc? Did you understand the content?
4. How does the way mental health and adverse childhood experiences being addressed in the game make you feel?
5. How did the game make you feel? Did you feel negative feelings at any point? When/how did you feel?
6. Did you feel the game was presented in a helpful way? Did you feel “safe” while playing? (if so, what helped?)
7. Did the game format make it easier or harder for you to reflect on mental health and difficult life experiences? Why?
8. Did the game make you feel “in control” of the space and the story? How?
9. What are some good and bad things about using this game to discuss adverse childhood experiences and mental health?
10. Did you resonate with the game?
11. Did you feel the game gave you appropriate information about: information privacy; what the game does and does not do; the positives and potential negative consequences of playing; who made the game?
12. Did the game connect you to supportive resources? How? What resources have you found helpful? Did you use any of them?
13. Did the game motivate you in a positive way?

**Supplementary Material 3.**

**eTable 2.** Additional characteristics of the sample by all participants who completed the baseline assessment (n=36) and participants who accessed the serious game (n=22)

| Characteristics | All participants (n=36) | Participants who accessed the game (n=22) |
|---|---|---|
| **Trans status, n (%)** | | |
| Yes | 5 (13.9) | 3 (13.6) |
| No | 30 (83.3) | 18 (81.8) |
| Questioning | 1 (2.8) | 1 (4.5) |
| **Born in the UK, n (%)** | | |
| Yes | 29 (80.6) | 18 (81.8) |
| No | 7 (19.4) | 4 (18.2) |
| **Neurodivergence, n (%)** | | |
| ADHD/ADD or Attention Differences | | |
| *Considered to have or experience* | 13 (36.1) | 8 (36.4) |
| *Received diagnosis* | 4 (11.1) | 2 (9.1) |
| Autistic / autism spectrum condition | | |
| *Considered to have or experience* | 6 (16.7) | 5 (22.7) |
| *Received diagnosis* | 7 (19.4) | 5 (22.7) |
| Demand avoidance | | |
| *Considered to have or experience* | 9 (25.0) | 5 (22.7) |
| *Received diagnosis* | 0 | 0 |
| Dyscalculia | | |
| *Considered to have or experience* | 3 (8.3) | 1 (4.5) |
| *Received diagnosis* | 1 (2.8) | 1 (4.5) |

| | | |
|---|---|---|
| Dyslexia | | |
| *Considered to have or experience* | 2 (5.6) | 1 (4.5) |
| *Received diagnosis* | 3 (8.3) | 0 |
| Dyspraxia | | |
| *Considered to have or experience* | 4 (11.1) | 3 (13.6) |
| *Received diagnosis* | 3 (8.3) | 2 (9.1) |
| Intellectual/Learning disability | | |
| *Considered to have or experience* | 5 (13.9) | 1 (4.5) |
| *Received diagnosis* | 2 (5.6) | 2 (9.1) |
| Selective/situational mutism | | |
| *Considered to have or experience* | 5 (13.9) | 3 (13.6) |
| *Received diagnosis* | 1 (2.8) | 1 (4.5) |
| Language impairment | | |
| *Considered to have or experience* | 2 (5.6) | 1 (4.5) |
| *Received diagnosis* | 0 | 0 |
| Tourette's Syndrome | | |
| *Considered to have or experience* | 1 (2.8) | 0 |
| *Received diagnosis* | 0 | 0 |
| None of the above | 12 (33.3) | 7 (31.8) |
| **Education status, n (%)** | | |
| School or Sixth Form College | 22 (61.1) | 11 (50.0) |
| Higher education (e.g., university) | 5 (13.9) | 5 (22.7) |
| Further education | 1 (2.8) | 0 |
| Not in education | 8 (22.2) | 6 (27.3) |
| **Employment status, n (%)** | | |
| Full-time | 2 (5.6) | 2 (9.1) |

| | | |
|---|---|---|
| Part-time | 9 (25.0) | 8 (36.4) |
| Zero-hour contract/holiday job | 2 (5.6) | 2 (9.1) |
| Not employed | 23 (63.9) | 10 (45.5) |
| **Parent/Caregiver status, n (%)** | | |
| More than one parent/caregiver | 24 (66.7) | 16 (72.7) |
| One parent/caregiver | 11 (30.6) | 6 (27.3) |
| No parent/caregiver | 1 (2.8) | 0 |
| **Foster/residential care status, n (%)** | | |
| No | 31 (86.1) | 21 (95.5) |
| Yes, in the past | 3 (8.3) | 1 (4.5) |
| Yes, at the moment | 1 (2.8) | 0 |
| Prefer not to say | 1 (2.8) | 0 |
| **Parent/Caregivers born in the UK, n (%)** | | |
| Yes, one parent/caregiver | 8 (22.2) | 4 (18.2) |
| Yes, both | 16 (44.4) | 10 (45.5) |
| No | 12 (33.3) | 8 (36.4) |
| **Main parent/caregiver education, n (%)**[a] | | |
| Attended secondary school | 2 (5.7) | 0 |
| Completed GCSEs | 6 (17.1) | 3 (13.6) |
| AS/A2 level | 3 (8.6) | 2 (9.1) |
| Technical/Vocational training | 3 (8.6) | 2 (9.1) |
| Bachelor's degree | 6 (17.1) | 6 (27.3) |
| Master's degree or advanced/PhD | 5 (14.3) | 3 (13.6) |
| Don't know or other | 10 (28.6) | 6 (27.3) |

| | | |
|---|---|---|
| **Main parent/caregiver employment, n (%)[a]** | | |
| Employed | 27 (77.1) | 16 (72.7) |
| Student | 2 (5.7) | 1 (4.5) |
| Does not have a job/not a student | 5 (14.3) | 5 (22.7) |
| Other | 1 (2.9) | 0 |
| **Mental wellbeing (SWEMWBS), M(SD)** | 22.28 (4.98) | 22.41 (5.26) |
| **PTSD symptoms (CRIES), M(SD)** | 23.28 (12.02) | 21.82 (13.28) |
| Intrusion symptom severity | 11.56 (6.76) | 11.27 (7.34) |
| Avoidance symptom severity | 11.72 (6.36) | 10.55 (6.84) |
| **Difficulties with Emotion Regulation (DERS-SF), M(SD)** | 49.58 (11.03)[b] | 49.00 (11.38)[c] |
| DERS-SF Awareness, M(SD) | 8.75 (3.31)[b] | 8.36 (3.17)[c] |
| DERS-SF Clarity, M(SD) | 7.50 (2.47)[b] | 7.55 (2.58)[c] |
| DERS-SF Goals, M(SD) | 10.75 (3.41)[b] | 11.09 (3.36)[c] |
| DERS-SF Impulsivity, M(SD) | 6.08 (2.50)[b] | 5.64 (2.06)[c] |
| DERS-SF Non-Acceptance, M(SD) | 8.17 (3.24)[b] | 8.18 (3.40)[c] |
| DERS-SF Strategies, M(SD) | 8.33 (3.31)[b] | 8.18 (3.43)[c] |

a One response was missing for **all participants**; total N = 35

b Measure completed by participants aged ≥18 years only (n = 12)

c Measure completed by participants aged ≥18 years only (n = 11)

**Supplementary Material 4.**

**eTable 3.** Acceptability evaluation for the ACE of Hearts game (n=20)

| Items | Value, n (%) |
|---|---|
| **Affective attitude** | |
| I like ACE of Hearts game | 19/20 (95.0) |
| I dislike ACE of Hearts game[a] | 1/19 (5.0) |
| I enjoy using ACE of Hearts game | 17/20 (85.0) |
| I hate using ACE of Hearts game[a] | 0/19 |
| **Burden** | |
| ACE of Hearts game is easy to use | 17/20 (85.0) |
| ACE of Hearts game is hard to use | 4/20 (20.0) |
| ACE of Hearts game is confusing | 9/20 (45.0) |
| ACE of Hearts game is simple | 14/20 (70.0) |
| **Ethicality** | |
| I care about mental health | 20/20 (100.0) |
| Mental health is important to me | 20/20 (100.0) |
| Mental health is unimportant to me | 0/20 |
| It’s good to care about mental health | 20/20 (100.0) |
| **Intervention coherence** | |
| I can show a friend how to use ACE of Hearts game | 17/20 (85.0) |
| I understand how to use all of the features in ACE of Hearts game | 11/20 (55.0) |
| I can show a friend how to use all of the features in ACE of Hearts game | 12/20 (60.0) |

| | |
|---|---|
| **Perceived effectiveness** | |
| ACE of Hearts game can help me think about my life experiences | 16/20 (80.0) |
| I can better understand difficult life experiences because of ACE of Hearts game[a] | 11/19 (55.0) |
| ACE of Hearts game can help me acknowledge hard experiences I have been through[a] | 16/19 (80.0) |
| I am better able to accept my own life experiences because of ACE of Hearts game[a] | 7/19 (35.0) |
| **Self-efficacy** | |
| I am confident that I could use ACE of Hearts game even if I'm really busy | 8/20 (40.0) |
| I am confident that I would use ACE of Hearts game even if I'm not reminded to do it | 13/20 (65.0) |
| I am confident that I could use ACE of Hearts game to help me if I'm really busy | 4/20 (20.0) |

a One response was missing; total n = 19.

**Supplementary Material 5.**

**eTable 4.** Pre-post mental health outcomes

| | **Baseline** (T0), mean (SD), (n=19) | **1-month follow-up** (T1), mean (SD), (n=19) | **3-month follow-up** (T2), mean (SD), (n=19) | **Pre-post change** (T1-T0), mean difference, (95% CI) | **Pre-post change** (T2-T0), mean difference (95% CI) |
|---|---|---|---|---|---|
| **Depression (PHQ-9/PHQ-C)**, (n=19) | 9.95 (6.64) | 8.95 (4.87) | 8.74 (5.10) | **-1.00** (-0.73, 2.73) | **-1.21** (-1.01, 3.43) |
| **Anxiety (GAD-7)**, (n=19) | 9.16 (5.63) | 7.84 (4.84) | 7.79 (4.45) | **-1.32** (-1.38, 4.01) | **-1.37** (-0.66, 3.40) |
| **Mental wellbeing (SWEMWBS)**, (n=19) | 22.63 (5.62) | 23.00 (5.77) | 23.53 (4.68) | **0.37** (-2.16, 1.42) | **0.89** (-2.82, 1.03) |
| **Impact of events (CRIES-8)**, (n=19) | 19.89 (13.26) | 23.84 (12.78) | 20.68 (13.53) | **3.95** (-10.46, 2.56) | **0.79** (-4.20, 2.62) |
| **Emotion regulation (DERS-SF)[a],** (n=9) | 47.78 (12.04) | 49.67 (11.53) | 43.33 (12.70) | **1.89** (-7.53, 3.75) | **-4.44** (-0.48, 9.37) |

Abbreviations: SD: Standard Deviation; CI: Confidence Interval
a DERS-SF completed by participants aged 18 years or older

**Supplementary Material 6.**

**eTable 5.** Characteristics of participants who completed a process evaluation interview (n=15)

| Characteristics | Participants who completed an interview (n=15) |
|---|---|
| **Age (years), *M(SD)*** | 18.80 (3.63) |
| **Age group, n (%)** | |
| 12-15 years | 3 (20.0) |
| 16-17 years | 3 (20.0) |
| 18-24 years | 9 (60.0) |
| **Gender, n (%)** | |
| Woman/girl | 6 (40.0) |
| Man/boy | 6 (40.0) |
| Non-binary | 2 (13.3) |
| Prefer not to say | 1 (6.7) |
| **Ethnic background, n (%)** | |
| Black, African, Caribbean or Black British | 2 (13.3) |
| Asian (various) or Asian British | 1 (6.7) |
| White (various) or white other | 11 (73.3) |
| Irish | 1 (6.7) |
| **Sexual orientation, n (%)** | |
| Heterosexual/straight | 3 (20.0) |
| Gay or Lesbian | 2 (13.3) |
| Bisexual or pansexual | 6 (40.0) |
| Asexual | 2 (13.3) |
| Don’t know | 2 (13.3) |

| | |
|---|---|
| **Neurodivergence, n (%)** | |
| None (of those listed) | 3 (21.4) |
| ADHD/ADD or Attention Differences | 6 (40.0) |
| Autistic / autism spectrum condition | 10 (66.7) |
| Other[a] | 6 (42.9) |
| **Neurodiversity diagnosis received, n (%)** | |
| None | 8 (53.3) |
| One diagnosis | 4 (26.7) |
| 2 diagnoses | 1 (6.7) |
| 3+ diagnoses | 2 (13.3) |
| **Geographical location, n (%)** | |
| City/urban area | 8 (53.3) |
| Small town/rural area | 6 (40.0) |
| By the sea/coastal area | 1 (6.7) |

a Includes category options: Demand Avoidance, Dyscalculia, Dyslexia, Dyspraxia, Intellectual Learning Disability, Selective or Situational Mutism, Language Impairment.
